# Modelling Under-Five Mortality in Zambia Using Advanced Survival Approaches: A Comparative Analysis of Pooled Demographic and Health Survey Data

**DOI:** 10.64898/2026.09.29.26364267

**Authors:** Samuel Mutasha, Given Moonga, Thelma Mwala, Kwame Shanaube

## Abstract

**Background:** Under-five mortality remains a major public health challenge in Zambia despite substantial reductions over recent decades. Demographic and Health Survey (DHS) data are commonly analysed using Cox proportional hazards models; however, violations of the proportional hazards assumption and unobserved cluster-level heterogeneity may compromise inference. This study compared advanced survival modelling approaches to determine the most appropriate framework for analysing under-five mortality using pooled Zambia DHS data.

**Methods:** We conducted a pooled secondary analysis of the 2018 and 2024 Zambia Demographic and Health Survey child recode datasets. The analysis included 9,982 children, among whom 266 died before their fifth birthday. A survey-weighted Cox proportional hazards model was fitted as the reference model and assessed using Schoenfeld residual diagnostics. To address identified methodological limitations, an extended Cox model, a shared-frailty model, and a Bayesian survival model were subsequently fitted. Model performance, assumptions, and the consistency of adjusted hazard ratio estimates were compared across modelling approaches.

**Results:** The global Schoenfeld test indicated violation of the proportional hazards assumption (χ²=51.26, p=0.002). In the adjusted extended Cox model, male children had higher mortality risk than females (aHR 1.95, 95% CI 1.40–2.73), while children with gestation ≥9 months had lower mortality risk than those with shorter gestation (aHR 0.21, 95% CI 0.14–0.32). Average and large birth size were associated with lower mortality compared with small birth size (aHR 0.47, 95% CI 0.34–0.66 and aHR 0.57, 95% CI 0.39–0.84, respectively). Newborn health checks before discharge were associated with lower mortality (aHR 0.50, 95% CI 0.38–0.66), as was any antenatal care (aHR 0.25, 95% CI 0.09–0.75). A high-risk fertility pattern involving fourth or higher births with an interval ≥24 months was also associated with lower mortality (aHR 0.57, 95% CI 0.41–0.80). The shared-frailty model produced similar estimates, including increased mortality among male children (aHR 1.39, 95% CI 1.08–1.78) and twins or multiple births (aHR 2.74, 95% CI 1.36–5.53). Bayesian estimates were broadly consistent with the extended Cox model.

**Conclusions:** Advanced survival models provided a more robust analytical framework than the conventional Cox model for evaluating under-five mortality in nationally representative DHS data. While the extended Cox model accommodated time-varying effects, the shared-frailty model captured unobserved cluster-level heterogeneity, and the Bayesian model confirmed the stability of parameter estimates. Incorporating these complementary approaches can improve the validity of survival analyses and strengthen evidence for child survival research and policy in low- and middle-income settings.

## Introduction

Under-five mortality has declined substantially worldwide over the past three decades, reflecting major improvements in maternal and child health, vaccination, nutrition, and access to essential health services. Globally, the under-five mortality rate declined from approximately 93 deaths per 1,000 live births in 1990 to 38 deaths per 1,000 live births in 2019, representing a reduction of almost 60%^1,2^. During the same period, the annual number of under-five deaths declined substantially, although millions of children continue to die before reaching their fifth birthday^1–3^. Progress in neonatal survival has been comparatively slower. Neonatal mortality declined from approximately 37 deaths per 1,000 live births in 1990 to about 18 per 1,000 live births by 2017–2019, resulting in neonatal deaths accounting for an increasing proportion of under-five deaths^2–5^. These trends indicate that, despite substantial global progress, preventable child mortality remains an important public health challenge, particularly during the neonatal period.

The burden is disproportionately concentrated in sub-Saharan Africa (SSA), which continues to have the highest under-five mortality rates globally. Around 2015, under-five mortality varied considerably across SSA countries, ranging from approximately 16 to 135 deaths per 1,000 live births^5–7^. In 2019, mortality in the African region remained substantially above the Sustainable Development Goal (SDG) target of 25 deaths per 1,000 live births, and many countries, particularly in West and Central Africa, remain off track to achieve SDG 3.2 without accelerated progress^2,3,8,9^. Although several countries have achieved considerable reductions in child mortality, progress has been uneven both between and within countries. Subnational differences in socioeconomic conditions, access to healthcare, maternal characteristics, and living environments continue to produce marked inequalities in child survival ^6,8,10,11^.

Zambia has also experienced substantial improvements in child survival, although mortality remains high relative to global targets. Analyses of Zambia Demographic and Health Survey (ZDHS) data have demonstrated a marked decline in under-five mortality over the past two decades, from approximately 119 deaths per 1,000 live births in the late 2000s to around 60–65 deaths per 1,000 live births by 2018 ^8,9^. Despite this progress, the pace of decline has been uneven across the different stages of childhood. Neonatal mortality has declined more slowly than post-neonatal and childhood mortality, leaving newborn survival as an important remaining challenge ^2,4,8^. Furthermore, substantial socioeconomic and geographic inequalities persist, with higher mortality reported among children from poorer households, those born to mothers with lower educational attainment, and populations living in high-burden areas ^9^. Previous studies using ZDHS data have identified several determinants of under-five mortality, including male sex, multiple births, low birth weight or prematurity, short birth intervals, inadequate antenatal care, and maternal socioeconomic disadvantage ^9,12–14^.

Understanding these determinants requires analytical approaches that appropriately account for the time-to-event nature of child mortality and the hierarchical structure of survey data. Most DHS-based studies in Zambia and other low- and middle-income countries have used logistic regression or conventional Cox proportional hazards models to examine mortality determinants ^12–15^. Logistic regression treats mortality as a binary outcome and therefore does not account for when death occurs, while conventional Cox proportional hazards models assume that the effects of covariates remain constant over follow-up time. These assumptions may not adequately reflect the dynamic nature of mortality risk during the first five years of life, when the risk of death changes substantially with age. In addition, children sampled within the same household, community, or survey cluster may share environmental, socioeconomic, and healthcare characteristics, resulting in correlated outcomes that conventional models may not fully capture ^12,13,16^.

Advanced survival modelling approaches can address these limitations. Extended Cox models allow hazard ratios to vary over time when the proportional hazards assumption is violated, thereby capturing age-dependent effects of risk factors¹⁵˒¹⁷. Shared-frailty models account for unobserved heterogeneity and clustering of mortality risk within households, communities, or survey clusters ^12,13,17^. Bayesian survival models provide a flexible framework for incorporating uncertainty and estimating complex relationships, particularly when events are relatively uncommon or models involve multiple sources of variation ^15,18^. Together, these approaches provide complementary perspectives on the timing, magnitude, and contextual variation of child mortality risk.

Despite increasing use of advanced survival methods in global and African child-survival research, their application to nationally representative Zambian data remains limited. There is therefore a need to determine whether conclusions about the determinants of under-five mortality are robust across alternative modelling frameworks and whether more flexible approaches reveal important temporal or contextual patterns that may be missed by conventional methods.

This study therefore aimed to evaluate and compare four survival modelling frameworks a survey-weighted Cox proportional hazards model, extended Cox model, Bayesian survival model, and shared-frailty model for analysing pooled ZDHS data on under-five mortality. A secondary objective was to examine determinants of under-five mortality and assess the consistency and variability of predictor estimates across these complementary modelling approaches.

## Materials and methods

### Study Design and Data Source

This study was a pooled secondary analysis of nationally representative child-level data from the Zambia Demographic and Health Surveys conducted in 2018 and 2024^9,19^. The ZDHS uses a stratified two-stage cluster sampling design ^9,19^. In the first stage, enumeration areas are selected as clusters within strata defined by province and urban–rural residence, and in the second stage households are sampled from listings within each selected cluster ^9,20^. The DHS is nationally representative and designed to estimate demographic and health indicators across geographic areas. The child recode (KR) datasets draw on women aged 15–49 years and include complete birth histories and child survival information for all live births recorded in the surveys.

The two survey datasets were appended to create a pooled child-level analytical cohort. To preserve the hierarchical structure of the survey data across survey rounds, a survey-round indicator was retained, and a unique cluster identifier was generated by combining the survey year with the original DHS cluster number. The pooled survival-ready cohort included 18,632 eligible children, comprising 775 under-five deaths and 17,857 right-censored observations. After applying complete-case restrictions for all variables included in the multivariable models, the final analytical sample comprised 9,982 children, including 266 under-five deaths and 9,716 right-censored observations. A total of 8,650 children were excluded because of missing covariate information, among whom 509 deaths occurred. The participant selection process is summarized in Figure 1.

**Figure 1.**
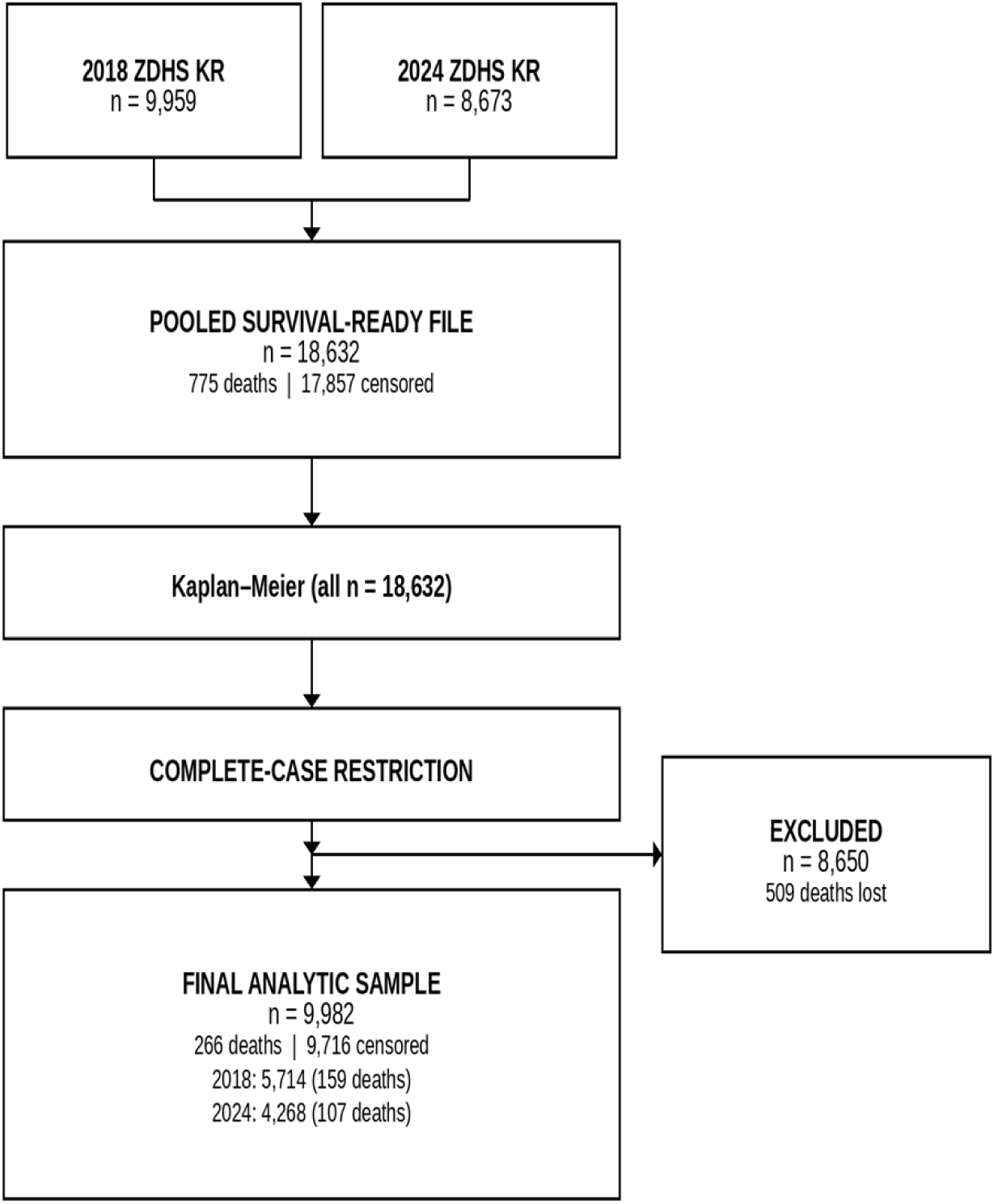
Selection workflow for the pooled ZDHS analytical cohort: 18,632 eligible children and 9,982 complete-case observations from the 2018 and 2024 ZDHS.

### Outcome and explanatory variables

The primary outcome was under-five mortality, defined as death before 60 completed months of age. Survival time was calculated from birth to death or censoring. For deceased children, age at death was derived from the DHS composite variable B6, converting days, months, and years to a common month scale; deaths on the day of birth were assigned a survival time of 1/60 month, a value below the smallest positive recorded death age, to avoid zero time bias Covariates were selected a priori based on epidemiological evidence and included child characteristics (sex, twin status, gestational age, birth size, and high-risk fertility pattern), maternal factors (education, pregnancy wantedness and smoking) antenatal care and obstetric characteristics (any antenatal care, malaria prophylaxis, tetanus toxoid injection, intestinal parasite treatment during pregnancy and mode of delivery), postnatal newborn health check before discharge (M74/M75, analysed as a time-varying covariate), and household characteristics (wealth index, improved water source, place of residence, and survey year). Tetanus toxoid injection and intestinal parasite treatment were assessed as candidate covariates but excluded from the final multivariable analyses due to substantial missingness. Postnatal newborn health before discharge was harmonized across survey rounds using M74 (check before discharge) and M75 (timing); M70/M71 were not pooled because question wording differs between 2018 and 2024. The newborn check indicator was entered as a time-varying covariate using a counting-process formulation.

### Statistical analysis

All analyses accounted for the DHS complex survey design, including sampling weights, stratification, and primary sampling units. Sampling weights were normalised before pooled analysis.

Time-varying newborn health check before discharge was implemented using a counting-process formulation with the R survival package tmerge() and tdc(), splitting follow-up at the recorded time of newborn health check before discharge for children who received the check. Proportional hazards were assessed with scaled Schoenfeld residuals (cox.zph). Where the PH assumption was violated, time-varying coefficients were modelled using (i) survSplit() step functions at 1 and 12 months with covariate:strata(tgroup) interactions, and (ii) parametric tt() terms with log(t + 1 month). A survey-weighted Cox model on the counting-process data served as the reference analysis, followed by extended Cox, shared-frailty, and Bayesian approaches as complementary frameworks.

Kaplan–Meier survival probabilities were estimated as:

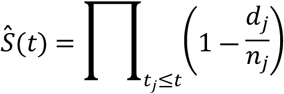

where *S*(*t*)represents the estimated probability of survival beyond time *t*, *d_j_*denotes the number of deaths occurring at event time *t_j_*, and *n_j_*represents the number of individuals at risk immediately prior to time *t_j_*. Kaplan–Meier estimators were used to describe the survival experience over time. The resulting survival curves were presented descriptively, while inferential conclusions were based on the regression models described below.

### Survey-weighted Cox proportional-hazards model

The survey-weighted Cox model estimated adjusted hazard ratios (HRs) with 95% confidence intervals while accounting for sampling weights, stratification, and clustering:

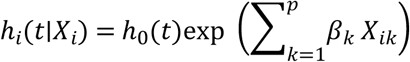

where ℎ*_i_*(*t*∣*X_i_*) denotes the hazard function for individual *i* at time *t*, ℎ_0_(*t*) represents the baseline hazard function, *X_ik_* is the value of the *k^t^*^ℎ^ covariate for individual *i*, and *β_k_* is the corresponding regression coefficient. The exponentiated coefficient, exp (*β_k_*), represents the hazard ratio associated with a one unit increase in covariate *k*, holding all other variables constant.

### Extended Cox modelling

Covariates with significant Schoenfeld tests (Table 2) were entered into an extended Cox model with tt() terms using the transformation log(t + 1 month). The resulting model included both the main effect and its interaction with log(t + 1 month) (Table 3). Supplementary Table S compares univariable, multivariable PH, and extended time-dependent estimates (Hellmund et al., Cancers 2020).

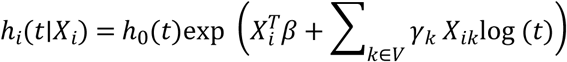

where ℎ*_i_*(*t*∣*X_i_*) denotes the hazard function for individual *i* at time *t*, ℎ_0_(*t*)represents the baseline hazard function, *X^T^β* captures the standard proportional covariate effects, and *V* denotes the set of covariates that violated the proportional hazards assumption. The parameter *γ_k_* quantifies the extent to which the effect of covariate *k* changes as a function of log time.

### Bayesian survival and shared-frailty models

A Bayesian survival model was fitted using **brms** in R with Hamiltonian Monte Carlo and the No-U-Turn Sampler. Weakly informative priors were specified, and posterior hazard ratios with 95% credible intervals were reported. Convergence was assessed using trace plots, effective sample sizes, and Gelman–Rubin statistics. A gamma-distributed shared-frailty model was used to account for unobserved cluster-level heterogeneity:

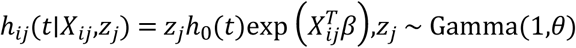

where ℎ*_ij_*(*t*∣*X_ij_*,*z_j_*) denotes the hazard function for individual *i* within cluster *j*, ℎ_0_(*t*) represents the baseline hazard function, *X^T^β* denotes the fixed covariate effects, and *z_j_*is the cluster specific multiplicative frailty term assumed to follow a gamma distribution with mean 1 and variance *θ*.

### Software

Analyses were conducted using Stata 17 for survey-weighted procedures and R version 4.5 for survival modelling, diagnostics, and Bayesian analysis. This dual-platform approach enhanced reproducibility and analytical flexibility.

## Results

### Descriptive survival patterns

Kaplan–Meier survival curves were used to describe differences in under-five survival across selected child and household characteristics before multivariable modelling (Figure 2). Survival differences were observed across sex, household wealth, twin status, and gestational age categories, with the greatest early-life separation observed among children with multiple births and those born at shorter gestational ages. These findings provided descriptive evidence of heterogeneity in survival experience and informed subsequent multivariable survival analyses.

**Figure 2.**
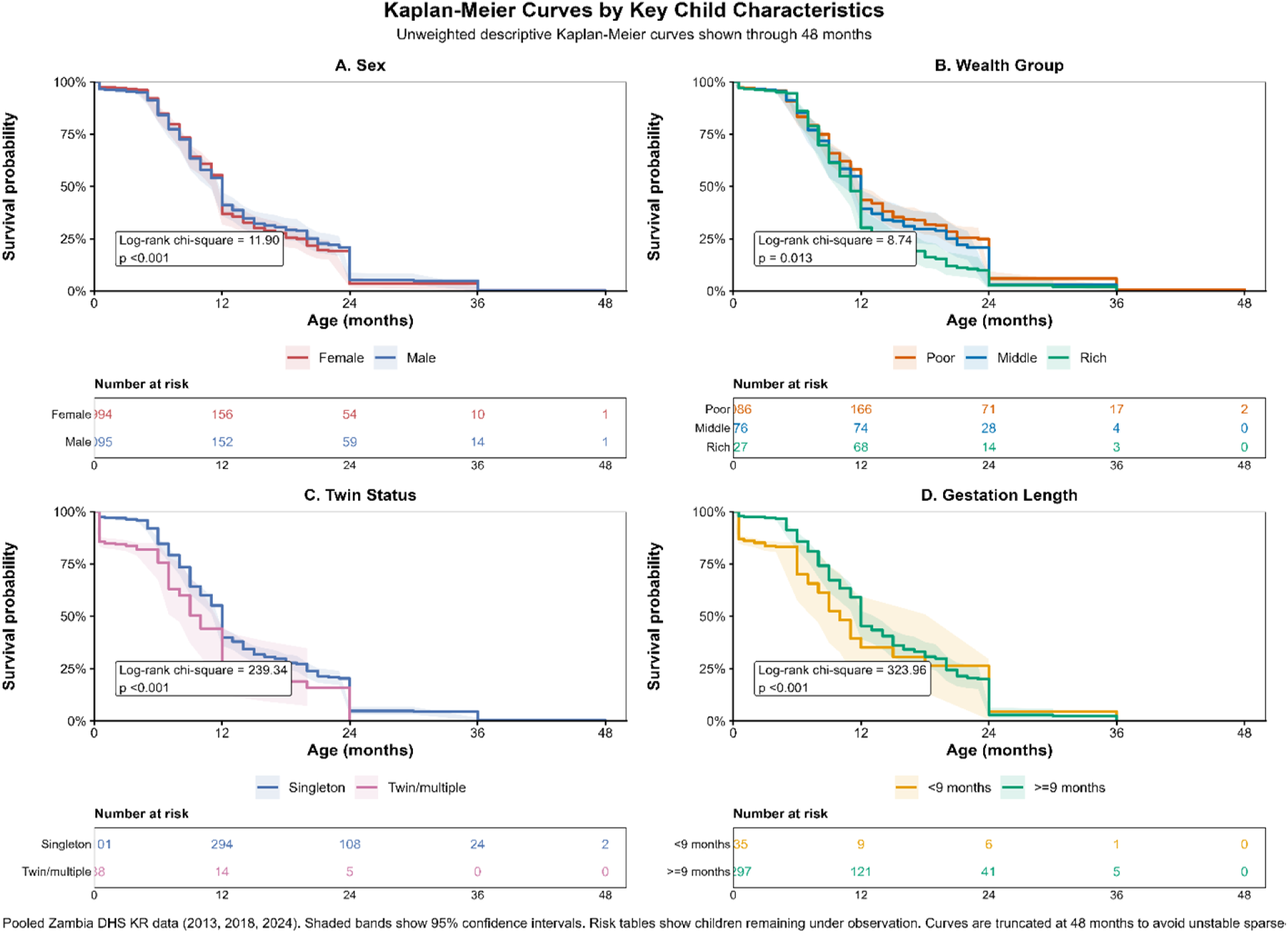
Kaplan–Meier survival experience demonstrating heterogeneity before multivariable survival modelling.

### Reference survey-weighted Cox model

The survey-weighted Cox proportional hazards model provided the initial design-adjusted reference analysis (Table 1). Several biological and early-life factors showed strong associations with under-five mortality. Male children had higher mortality risk compared with females (HR 1.40 (1.10–1.79); p=0.022), while twin or multiple births were associated with increased mortality (HR 2.50 (1.28–4.86); p=0.059). Gestation ≥9 months was strongly protective compared with shorter gestation (HR 0.31 (0.22–0.42); p<0.001). Newborn health checks before discharge, modelled as a time-varying covariate using M74/M75, were associated with lower mortality risk after the check occurred (HR 0.50 (0.37–0.66); p<0.001). Any antenatal care was associated with lower mortality (HR 0.26 (0.09–0.78); p=0.018), whereas most household socioeconomic indicators and several other maternal healthcare variables showed limited evidence of association after adjustment. However, this model was considered a reference analysis only because the proportional hazards assumption was not satisfied.

**Table 1.** Baseline survey-weighted Cox proportional-hazards model for the reference analysis of under-five mortality (n = 9,982; deaths = 266)

| Variable | HR (95% CI) | p-value |
| --- | --- | --- |
| **A. Child-Related Factors** |  |  |
| **Child sex** |  |  |
| Female | Reference | — |
| Male | 1.40 (1.10–1.79) | 0.023 |
| **Multiple birth status** |  |  |
| Singleton | Reference | — |
| Twin/multiple birth | 2.53 (1.30–4.92) | 0.055 |
| **Gestational age at birth** |  |  |
| <9 months | Reference | — |
| $\geq 9$ months | 0.31 (0.22–0.42) | <0.001 |
| **Child size at birth** |  |  |
| Small | Reference | — |
| Average | 0.46 (0.33–0.64) | <0.001 |
| Large | 0.56 (0.38–0.82) | 0.012 |
| **High-risk fertility pattern** |  |  |
| First birth | Reference | — |
| 2nd–3rd birth, <24-month interval | 1.27 (0.74–2.20) | 0.372 |
| 2nd–3rd birth, ≥24-month interval | 0.75 (0.55–1.03) | 0.135 |
| ≥4th birth, <24-month interval | 1.20 (0.71–2.03) | 0.531 |
| ≥4th birth, ≥24-month interval | 0.58 (0.41–0.81) | 0.005 |
| **B. Maternal Factors** |  |  |
| **Maternal education** |  |  |
| No education | Reference | — |
| Primary | 1.73 (0.97–3.08) | 0.054 |
| Secondary | 1.48 (0.80–2.74) | 0.212 |
| Higher | 1.87 (0.85–4.11) | 0.142 |
| **Maternal smoking** |  |  |
| Non-smoker | Reference | — |
| Smoker | 0.83 (0.17–3.96) | 0.859 |
| **Pregnancy wantedness** |  |  |
| Unwanted/mistimed | Reference | — |
| Wanted | 1.09 (0.85–1.40) | 0.542 |
| **C. Antenatal and Obstetric Factors** |  |  |
| **Any antenatal care (ANC)** |  |  |
| No | Reference | — |
| Yes | 0.26 (0.09–0.76) | 0.016 |
| **Malaria prophylaxis during pregnancy (SP/IPT)** |  |  |
| No | Reference | — |
| Yes | 0.98 (0.55–1.75) | 0.958 |
| **Tetanus toxoid injections** |  |  |
| None | Reference | — |
| Once | 0.98 (0.70–1.38) | 0.933 |
| 2–7 times | 1.12 (0.78–1.62) | 0.570 |
| **Intestinal parasite treatment during pregnancy** |  |  |
| No | Reference | — |
| Yes | 1.21 (0.90–1.62) | 0.256 |
| **Mode of delivery** |  |  |
| Vaginal/other | Reference | — |
| Caesarean section | 1.29 (0.84–1.96) | 0.324 |
| **D. Postnatal and Residential Factors** |  |  |
| **Newborn health check before discharge (time-varying)** |  |  |
| No check before discharge | Reference | — |
| Check before discharge | 0.49 (0.37–0.65) | <0.001 |
| **Place of residence** |  |  |
| Rural | Reference | — |
| Urban | 0.77 (0.53–1.10) | 0.147 |
| **Survey year** |  |  |
| 2018 | Reference | — |
| 2024 | 1.03 (0.80–1.33) | 0.838 |
| **E. Socioeconomic Factors** |  |  |
| **Household wealth index** |  |  |
| Poor | Reference | — |
| Middle | 1.13 (0.80–1.59) | 0.566 |
| Rich | 1.13 (0.73–1.75) | 0.585 |
| **Improved water source** |  |  |
| Unimproved | Reference | — |
| Improved | 0.94 (0.70–1.25) | 0.694 |

HR = Hazard ratio; CI = 95% confidence interval; Ref = Reference category; SP/IPT = Sulfadoxine-pyrimethamine / intermittent preventive treatment.

### Assessment of proportional hazards assumptions

The global Schoenfeld residual test demonstrated violation of the proportional hazards assumption (χ²(26)=51.26; p=0.002), indicating that the conventional Cox model was insufficient as the final analytical approach. Covariate-specific violations were identified for child sex, gestational age, maternal education, and caesarean delivery (Table 2). Diagnostic scaled Schoenfeld plots (Figure 3) and parametric tt() residual plots with fitted reference lines (Figure 4) supported incorporation of time-dependent effects in the extended Cox model.

**Figure 3.**
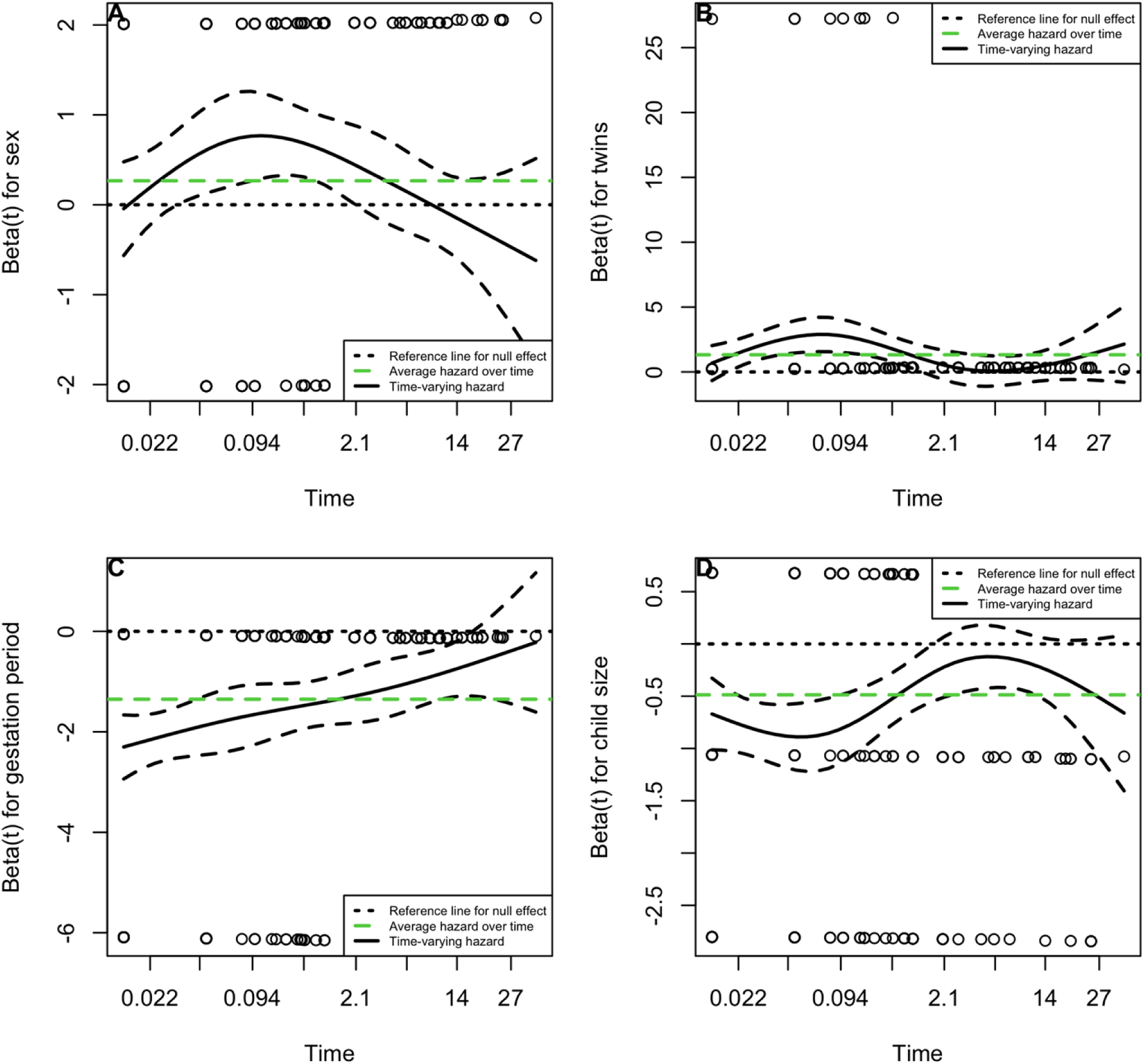
The effect of selected covariates on under-five mortality varies over time. The horizontal time axis is in km transformed scale, which is the default setting in the cox.zph() function. The dashed lines are the lower and upper limits of the confidence interval for each covariate effect. Panels show sex (top-left), twin status (top-right), gestation period (bottom-left), and child size (bottom-right).

**Figure 4.**
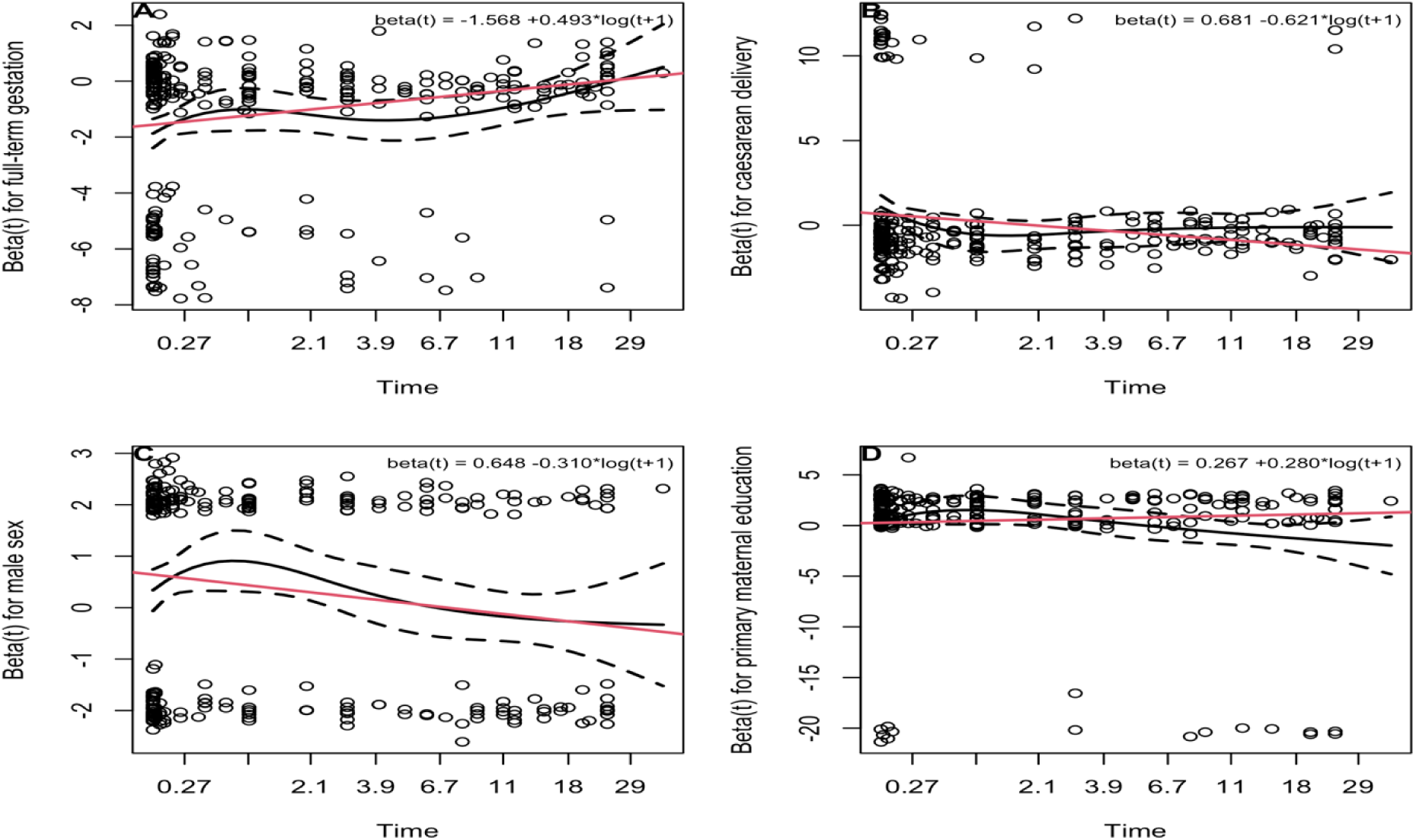
Parametric time-varying coefficient fits for covariates that violated proportional hazards in the multivariable reference model. The time axis was transformed as log(t + 1 month) to match the tt() specification; the black trace shows the scaled Schoenfeld-based coefficient and dashed lines its confidence interval. The red line shows the fitted parametric coefficient beta(t). Panels show Gestation ≥9 months (top-left), caesarean delivery (top-right), male sex (bottom-left), and primary maternal education (bottom-right).

**Table 2.** Assessment of proportional-hazards assumptions supporting the need for extended Cox modelling.

| Variable | ρ (Schoenfeld) | χ<sup>2</sup> | df | p-value | PH violated? |
| --- | --- | --- | --- | --- | --- |
| Child sex (Male vs. Female) | +7.641 | 1.00 | 1 | 0.006 | Yes |
| Multiple birth (twin) | +3.187 | 1.00 | 1 | 0.074 | No |
| Gestational age (≥9 months) | +8.620 | 1.00 | 1 | 0.003 | Yes |
| Child size at birth | +1.524 | 2.00 | 2 | 0.467 | No |
| Maternal education | +11.433 | 3.00 | 3 | 0.010 | Yes |
| Maternal smoking | +1.904 | 1.00 | 1 | 0.168 | No |
| Pregnancy wantedness | +0.171 | 1.00 | 1 | 0.679 | No |
| Any antenatal care | +0.192 | 1.00 | 1 | 0.661 | No |
| Malaria prophylaxis (SP/IPT) | +0.537 | 1.00 | 1 | 0.464 | No |
| Tetanus toxoid injections | +3.962 | 2.00 | 2 | 0.138 | No |
| Intestinal parasite treatment | +0.355 | 1.00 | 1 | 0.551 | No |
| Caesarean section | +5.729 | 1.00 | 1 | 0.017 | Yes |
| Newborn health check before discharge | +2.694 | 1.00 | 1 | 0.101 | No |
| Place of residence (Urban vs. Rural) | +0.000 | 1.00 | 1 | 0.999 | No |
| Household wealth index | +0.370 | 2.00 | 2 | 0.831 | No |
| High-risk fertility pattern | +2.169 | 4.00 | 4 | 0.705 | No |
| Improved water source | +0.551 | 1.00 | 1 | 0.458 | No |
| Survey year (2024 vs. 2018) | +0.019 | 1.00 | 1 | 0.891 | No |
| GLOBAL |  | 51.26 | 26 | 0.002 | Yes |

Note. *Test conducted on the cluster-robust Cox model (vce(cluster cluster\_uid); estat phtest, detail). ρ* *= Pearson correlation of scaled Schoenfeld residuals with analysis time.*

Figure 3 displays univariable diagnostic panels for selected covariates, whereas Figure 4 overlays multivariable Schoenfeld residuals with parametric tt() reference lines for covariates that violated proportional hazards in the adjusted reference model.

Parametric tt() fits in Figure 4 quantified how hazard ratios evolved over follow-up for covariates that violated proportional hazards in the multivariable reference model. The association between gestation ≥9 months and mortality was strongest in the first month (HR 0.29) and weakened by 60 months (HR 1.58) Caesarean delivery was associated with higher apparent hazard shortly after birth (HR 1.28 at 1 month) but substantially lower hazard thereafter (HR 0.15 at 60 months), consistent with confounding by acute obstetric indication rather than a sustained harmful effect. Male sex conferred the greatest excess mortality in the first month (HR 1.54), with the association attenuating over time (HR 0.53 at 60 months). Compared with no formal education, primary maternal education showed a modest early association (HR 1.59 at 1 month) that strengthened with longer follow-up (HR 4.13 at 60 months), reflecting progressively diverging survival patterns.

### Extended Cox model with time-dependent effects

The extended Cox model accounted for covariates whose effects changed over follow-up time and was considered the primary frequentist model. The direction and magnitude of major associations remained consistent with the reference model (Table 3). Male sex remained associated with increased mortality risk (HR 1.38; 95% CI: 1.08–1.76; p=0.028), while Gestation ≥9 months (HR 0.38; 95% CI: 0.26–0.54; p<0.001), average birth size (HR 0.45; 95% CI: 0.33–0.63; p<0.001), large birth size (HR 0.55; 95% CI: 0.37–0.81; p=0.010), newborn health check before discharge (HR 0.62; 95% CI: 0.45–0.86; p=0.015), and any antenatal care (HR 0.26; 95% CI: 0.09–0.78; p=0.018) remained associated with reduced mortality risk.

**Table 3.** Comparison of hazard estimates across advanced survival modelling approaches for under-five mortality.

| Variable | Extended Cox HR (95% CI) | Bayesian Survival HR (95% CrI) | Multilevel Survival HR (95% CI) |
| --- | --- | --- | --- |
| **A. Child-Related Factors** |  |  |  |
| **\*Child sex** |  |  |  |
| Female | 1.00 (Ref) | 1.00 (Ref) | 1.00 (Ref) |
| Male | 1.95 (1.40–2.73); p<0.001 | 1.95 (1.34–2.85) | 1.39 (1.08–1.78); p=0.010 |
| *Time interaction: male x log(time + 1 month)* | *0.71 (0.57–0.89)* | — | — |
| **Twin status** |  |  |  |
| Singleton | 1.00 (Ref) | 1.00 (Ref) | 1.00 (Ref) |
| Twin/multiple | 2.51 (1.29–4.88); p=0.058 | 2.53 (1.01–6.27) | 2.74 (1.36–5.53); p=0.005 |
| **\*Gestation length** |  |  |  |
| <9 months | 1.00 (Ref) | 1.00 (Ref) | 1.00 (Ref) |
| ≥9 months | 0.21 (0.14–0.32); p<0.001 | 0.21 (0.14–0.32) | 0.30 (0.22–0.42); p<0.001 |
| *Time interaction: ≥9 months x log(time + 1 month)* | *1.59 (1.13–2.24)* | — | — |
| **Birth size** |  |  |  |
| Small | 1.00 (Ref) | 1.00 (Ref) | 1.00 (Ref) |
| Average | 0.47 (0.34–0.66); p<0.001 | 0.47 (0.32–0.71) | 0.46 (0.33–0.66); p<0.001 |
| Large | 0.57 (0.39–0.84); p=0.015 | 0.57 (0.36–0.90) | 0.58 (0.39–0.86); p=0.007 |
| **High-risk fertility** |  |  |  |
| First birth | 1.00 (Ref) | 1.00 (Ref) | 1.00 (Ref) |
| 2nd–3rd, <24 mo | 1.24 (0.72–2.14); p=0.429 | 1.24 (0.72–2.10) | 1.35 (0.77–2.37); p=0.295 |
| 2nd–3rd, ≥24 mo | 0.75 (0.55–1.02); p=0.130 | 0.75 (0.52–1.10) | 0.74 (0.54–1.02); p=0.063 |
| ≥4th, <24 mo | 1.18 (0.70–1.99); p=0.572 | 1.18 (0.68–2.10) | 1.18 (0.69–2.02); p=0.543 |
| ≥4th, ≥24 mo | 0.57 (0.41–0.80); p=0.004 | 0.57 (0.40–0.84) | 0.56 (0.39–0.79); p<0.001 |
| **B. Maternal Factors** |  |  |  |
| **\*Maternal education** |  |  |  |
| No education | 1.00 (Ref) | 1.00 (Ref) | 1.00 (Ref) |
| Primary | 1.36 (0.73–2.53); p=0.309 | 1.38 (0.76–2.49) | 1.68 (0.93–3.05); p=0.085 |
| *Time interaction: primary x log(time + 1 month)* | *1.27 (1.01–1.58)* | — | — |
| Secondary | 1.47 (0.79–2.71); p=0.224 | 1.48 (0.80–2.77) | 1.48 (0.79–2.77); p=0.223 |
| Higher | 1.82 (0.83–4.01); p=0.157 | 1.85 (0.82–4.24) | 1.82 (0.81–4.09); p=0.150 |
| **Maternal smoking** |  |  |  |
| Non-smoker | 1.00 (Ref) | 1.00 (Ref) | 1.00 (Ref) |
| Smoker | 0.84 (0.18–4.00); p=0.866 | 0.83 (0.10–6.56) | 0.84 (0.17–4.13); p=0.828 |
| **Pregnancy wantedness** |  |  |  |
| Unwanted/mistimed | 1.00 (Ref) | 1.00 (Ref) | 1.00 (Ref) |
| Wanted | 1.08 (0.84–1.39); p=0.578 | 1.08 (0.82–1.42) | 1.10 (0.85–1.42); p=0.477 |
| **C. Antenatal/Obstetric Factors** |  |  |  |
| **Any ANC** |  |  |  |
| No | 1.00 (Ref) | 1.00 (Ref) | 1.00 (Ref) |
| Yes | 0.25 (0.09–0.75); p=0.014 | 0.26 (0.09–0.76) | 0.21 (0.07–0.65); p=0.007 |
| **Malaria prophylaxis** |  |  |  |
| No | 1.00 (Ref) | 1.00 (Ref) | 1.00 (Ref) |
| Yes | 1.00 (0.56–1.78); p=0.996 | 1.00 (0.55–1.81) | 0.94 (0.52–1.69); p=0.839 |
| **\*Caesarean section** |  |  |  |
| No | 1.00 (Ref) | 1.00 (Ref) | 1.00 (Ref) |
| Yes | 1.82 (1.10–3.01); p=0.072 | 1.82 (0.94–3.52) | 1.34 (0.87–2.06); p=0.190 |
| *Time interaction: caesarean delivery x log(time + 1 month)* | *0.59 (0.34–1.00)* | — | — |
| **D. Postnatal and Residential Factors** |  |  |  |
| **\*\*Newborn health check before discharge** |  |  |  |
| No | 1.00 (Ref) | 1.00 (Ref) | 1.00 (Ref) |
| Yes (time-varying) | 0.50 (0.38–0.66); p<0.001 | 0.50 (0.36–0.70) | 0.48 (0.35–0.64); p<0.001 |
| **Place of residence** |  |  |  |
| Rural | 1.00 (Ref) | 1.00 (Ref) | 1.00 (Ref) |
| Urban | 0.78 (0.54–1.12); p=0.170 | 0.78 (0.54–1.11) | 0.78 (0.53–1.16); p=0.228 |
| **Survey year** |  |  |  |
| 2018 | 1.00 (Ref) | 1.00 (Ref) | 1.00 (Ref) |
| 2024 | 1.03 (0.80–1.33); p=0.840 | 1.03 (0.77–1.38) | 1.02 (0.77–1.35); p=0.896 |
| **Household wealth** |  |  |  |
| Poor | 1.00 (Ref) | 1.00 (Ref) | 1.00 (Ref) |
| Middle | 1.11 (0.79–1.57); p=0.610 | 1.11 (0.74–1.69) | 1.15 (0.81–1.64); p=0.427 |
| Rich | 1.12 (0.72–1.73); p=0.623 | 1.11 (0.72–1.74) | 1.04 (0.66–1.65); p=0.856 |
| **Improved water source** |  |  |  |
| Unimproved | 1.00 (Ref) | 1.00 (Ref) | 1.00 (Ref) |
| Improved | 0.94 (0.70–1.25); p=0.697 | 0.93 (0.67–1.30) | 0.92 (0.68–1.25); p=0.593 |

*HR = Hazard ratio; CI = 95% confidence interval; CrI = 95% credible interval (Bayesian model); Ref = Reference category. Bold*
*italic rows denote conceptual domain subheadings. p-values are reported for Cox-PH and multilevel models; Bayesian estimates*
*are summarised by posterior credible intervals only. p-values <0.05 indicate statistical significance. \*For covariates violating the*
*proportional hazard assumption, time-interaction estimates are included in the extended cox model. \*\*Newborn healthcheck was*
*before discharge was modelled as a time-varying covariate.*

### Bayesian survival model

The Bayesian survival model produced estimates comparable in direction and magnitude to the extended Cox model (Table 3). Posterior hazard ratios demonstrated consistent associations for key predictors, including male sex, gestational age, birth size, and newborn health check before discharge. The agreement between frequentist and Bayesian estimates suggests that the main conclusions were robust to alternative inferential frameworks.

### Shared-frailty model for cluster-level heterogeneity

The shared-frailty model was fitted to account for potential unmeasured clustering of mortality risk within DHS sampling clusters. Estimates were generally consistent with the extended Cox and Bayesian models. Male sex, full-term gestation, and newborn health check before discharge remained important predictors. Twin or multiple birth reached statistical significance in this model (HR 2.70; 95% CI: 1.33– 5.47; p=0.006), alongside male sex, full-term gestation, and newborn health check before discharge.

### Cross-model comparison: the principal methodological result

Comparison across the extended Cox, Bayesian, and shared-frailty models demonstrated substantial stability in the magnitude and direction of key associations (Table 3). The strongest and most consistent predictors of under-five mortality were male sex, gestational age, and newborn health checks before discharge. Several socioeconomic and service-coverage variables, including household wealth, place of residence, malaria prophylaxis, and tetanus vaccination, were not statistically significant after full adjustment across models.

## Discussion

This study evaluated the consistency of different survival modelling approaches for analysing under-five mortality using pooled 2018 and 2024 Zambia Demographic and Health Survey (ZDHS) data. Our findings demonstrate that key determinants of child survival remained broadly consistent across the extended Cox, shared-frailty, and Bayesian survival models, supporting the robustness of the observed associations. Although the survey-weighted Cox proportional hazards model remains widely used for analysing time-to-event outcomes, its validity depends on the proportional hazards assumption ^21^.

In our study, the global Schoenfeld residual test indicated violation of this assumption, supporting the use of the extended Cox model to account for time-varying covariate effects and improve model specification^17,22^. The shared-frailty model further addressed unobserved clustering of mortality risk within DHS sampling units, an important consideration in African DHS datasets where children are nested within households and communities^18,21,23^. Similarly, the Bayesian survival model provided an alternative probabilistic framework for assessing parameter stability, consistent with previous DHS analyses demonstrating general agreement between Bayesian and classical survival approaches while occasionally identifying additional predictors^24^. Collectively, these findings indicate that the observed associations were not dependent on a single analytical approach but remained consistent across complementary survival models.

Male children had a consistently higher risk of under-five mortality compared with females, a pattern widely reported across African survival studies and global child mortality analyses ^25–27^. This association is thought to reflect biological vulnerability among male infants and children, including differences in immune function, inflammatory responses, and susceptibility to infection-related complications ^28,29^. The persistence of this association across multiple modelling frameworks highlights the importance of sex-disaggregated monitoring of neonatal and child health outcomes to ensure that interventions adequately address differences in vulnerability. Twin and multiple births were associated with substantially increased mortality risk compared with singleton births, consistent with evidence from DHS studies across sub-Saharan Africa ^30,31^. The excess mortality associated with multiple births is often concentrated in the neonatal period, underscoring the need for intensive early newborn care for twins and higher-order multiple births ^15,32^. Multiple births are associated with increased risks of prematurity, low birth weight, neonatal complications, and feeding difficulties, which contribute to higher mortality in resource-limited settings^31^. Strengthening district-level newborn care capacity, including kangaroo mother care and essential newborn care services, remains a key strategy for improving outcomes among these high-risk infants ^33^.

Gestational age was among the strongest predictors of under-five mortality, with gestation ≥9 months showing a lower estimated mortality risk than <9 months in the reference model but with variation in the association over follow-up in the extended cox model.This finding aligns with extensive evidence identifying preterm birth as one of the leading causes of neonatal mortality globally, particularly in low- and middle-income countries^34,35^. The increased vulnerability among preterm infants is mediated through physiological immaturity, impaired host defence mechanisms, respiratory complications, and increased susceptibility to infection^15^. In Zambia, reducing mortality related to prematurity requires integrated maternal and newborn health strategies addressing upstream determinants such as maternal malaria, anaemia, adolescent pregnancy, and inadequate maternal nutrition.

Newborn health checks before discharge were among the most consistent protective factors identified in this study. When modelled as a time-varying covariate reflecting whether a newborn health check occurred before discharge, this exposure was associated with reduced mortality risk across all modelling approaches. These findings reinforce global evidence that the first days following birth represent a critical period for preventing neonatal and child deaths ^36^. Previous evidence from Zambia has also demonstrated that strengthening maternal and newborn health services with postnatal components can contribute to reductions in neonatal mortality^36^. Improving facility-based newborn assessment before discharge should therefore remain a central priority within child survival programmes ^37,38^.

The association between caesarean delivery and mortality should be interpreted cautiously. Although the proportional hazards assumption for caesarean delivery was violated, the adjusted association with mortality was not statistically significant in the reference or extended models. Any apparent relationship should therefore be interpreted as confounding by indication, whereby caesarean delivery is more common among pregnancies complicated by obstructed labour, haemorrhage, fetal distress, and other high-risk conditions associated with increased neonatal mortality^39^. Since DHS data do not distinguish elective from emergency caesarean procedures, causal interpretation remains limited. The implication is therefore not reduced access to caesarean delivery but improved obstetric referral systems, timely decision-making, and quality of perioperative maternal and neonatal care ^40^.

Maternal tobacco use was not statistically significant after full adjustment in the present analysis, although tobacco exposure remains an important upstream risk factor in the broader literature ^41,42^. Tobacco exposure has established adverse effects on pregnancy and child health, and its effects may interact with other environmental exposures, including household air pollution from solid fuels, which remains common in many rural communities in Zambia ^43^. These findings should nonetheless be interpreted cautiously given the small number of exposed children. Geographically targeted tobacco prevention and household air-quality interventions may provide greater benefit than uniform approaches alone.

Any antenatal care was associated with lower mortality in the adjusted models, whereas several other established determinants, including household wealth, place of residence, malaria prevention interventions, and tetanus vaccination, were not statistically significant after full adjustment. This finding should not be interpreted as evidence that these factors are unimportant. In multivariable survival models, correlated predictors, mediation pathways, and overadjustment can attenuate associations between upstream determinants and mortality outcomes ^44^. Poverty, maternal education, and healthcare access remain well-established structural determinants of child survival inequalities across sub-Saharan Africa. Addressing these underlying drivers remains essential for achieving SDG 3.2 targets and reducing inequities in child survival.

## Conclusion

This study demonstrates that advanced survival modelling approaches provide important methodological advantages for analysing DHS-based child mortality data. The violation of proportional hazards assumptions and the presence of cluster-level heterogeneity in pooled ZDHS data highlight the limitations of relying solely on conventional Cox regression models. The consistency of estimates across extended Cox, Bayesian, and shared-frailty models strengthens confidence in the identified epidemiological associations, particularly the increased mortality risk associated with male sex, multiple births, the lower estimated risk with gestation ≥9 months, and the protective effect of newborn health check before discharge. Beyond identifying determinants of child mortality, this study highlights the importance of selecting analytical approaches that appropriately reflect the time-dependent and hierarchical nature of child survival data. Continued investment in antenatal services, facility-based newborn checks before discharge, and early care for twins and preterm infants remains essential for achieving SDG 3.2 targets, alongside methodological improvements in future child survival research.

### Limitations

This study has several limitations. First, the retrospective nature of DHS data limits causal interpretation and may be affected by recall error, survivor bias, and potential misclassification of child survival outcomes. Second, several clinically important variables, including measured birth weight, causes of death, and the indication or timing of caesarean delivery, were unavailable, limiting deeper interpretation of some associations. Third, complete-case analysis reduced the analytic sample from 18,632 to 9,982 children, and exclusion of observations with missing covariates may have introduced selection bias if missingness was not random. Fourth, although the Bayesian model demonstrated satisfactory convergence and used weakly informative priors, posterior estimates may remain sensitive to prior specification. Finally, while the shared-frailty model accounted for unmeasured cluster-level variation, the absence of detailed community-level contextual variables in the DHS limited the ability to identify the specific sources of this heterogeneity.

## List of Abbreviations / Acronyms

AHR: Adjusted Hazard Ratio
CI: Confidence Interval
CrI: Credible Interval
DHS: Demographic and Health Survey
HMC: Hamiltonian Monte Carlo
HR: Hazard Ratio
LMICs: Low- and Middle-Income Countries
MCMC: Markov Chain Monte Carlo
PH: Proportional Hazards
PSU: Primary Sampling Unit
SDGs: Sustainable Development Goals
SSA: Sub-Saharan Africa
WHO: World Health Organization

## Acknowledgements

The authors gratefully acknowledge the Demographic and Health Surveys Program, the Government of Zambia, participating households, field data collectors, and survey administrators for their invaluable contributions to the 2018 and 2024 Zambia Demographic and Health Surveys, without which this analysis would not have been possible.

## Author Contributions

SM, TM led the conceptualisation of the study, performed all statistical analyses, and prepared the initial manuscript draft. GM and KS contributed to the conceptualisation of the study and provided overall oversight, methodological guidance, and critical review of both the analysis and manuscript. All authors reviewed the final manuscript and approved it for submission.

## Data Availability

The data analyzed in this study are derived from the 2018 and 2024 Zambia Demographic and Health Surveys, which are publicly available through the <u>Demographic and Health Surveys Program</u>. Access to these data is governed by DHS data access approval procedures, and researchers may submit a formal request through the DHS Program data repository portal.

## Declarations

### Ethics Approval and Consent to Participate

The 2018 and 2024 Zambia Demographic and Health Surveys were conducted in full accordance with the ethical principles set forth in the Declaration of Helsinki. Ethical approval was secured from the relevant national ethics authorities in Zambia, with technical and procedural oversight provided by the Demographic and Health Surveys Program. Written informed consent was obtained from all participants prior to the commencement of data collection.

The present study constitutes a secondary analysis of de-identified, publicly available DHS data, accessed through an approved DHS data request. As the analysis involved no direct participant contact and used only anonymised data, no additional ethical review or approval was required.

### Consent for Publication

Not applicable. This manuscript contains no individually identifiable data or personal information pertaining to any participant.

